# Sleep improvement for metabolic health: A feasibility trial of a digital sleep treatment in people with insomnia and non-diabetic hyperglycaemia

**DOI:** 10.1101/2025.10.27.25338890

**Authors:** Rachel Sharman, David Ray, Andrew Farmer, Poppy C.E. Green, Victoria Harris, Fredrik Karpe, Colin A. Espie, Diana Mantripp, Thomas Marjot, Niall M. McGowan, Jeremy W. Tomlinson, Simon D. Kyle

## Abstract

Insomnia may play a causal role in type 2 diabetes (T2D). Addressing insomnia through cognitive behavioural therapy (CBTi) in people with non-diabetic hyperglycaemia could potentially reduce the risk of progression to T2D. To inform a future randomised trial, we performed a feasibility study of digital CBT (dCBTi) in individuals at increased risk of T2D. Participants were identified from ten primary care practices in the UK and given access to dCBTi. Outcomes were evaluated at baseline (week-0) and post-treatment (week-11). Primary feasibility outcomes were ability to recruit and treatment engagement. We also quantified within-group mean change (95%CI) in insomnia severity (Insomnia Severity Index), health-related quality of life (EQ-5D-3L), depression (Center for Epidemiologic Studies Depression Scale), chronotype (reduced Morningness-Eveningness Questionnaire), sleep (7-day actigraphy and diary), continuous glucose monitoring (7-days) and fasting blood metabolites (insulin, lipids, glucose, and C-reactive protein). The recruitment target was 20. Of 242 people completing screening, 36 were eligible, and 24 were enrolled (age 65.5±12.4 years, 70.8% female). Twenty-three (96%) completed post-intervention assessments. Treatment engagement was excellent (83.3% completed ≥4 sessions). The intervention was associated with a large reduction in insomnia severity [−4.7(95%CI:−6.2 to −3.2), *d*=−1.4] and medium reduction in depressive symptoms [−2.7(95%CI:−5.1 to −0.2), *d*=−0.5]. Sleep diary parameters tended to show greater improvement following intervention relative to actigraphy. There was evidence of a reduction in serum lactate, glycerol and triglycerides but no clear change in glucose or insulin. Results suggest a full trial is likely feasible and that people with NDH find the intervention acceptable and beneficial.

**Trial Registration:** This trial was prospectively registered on the UKs clinical study registry, the ISRCTN (ISRCTN19682964, https://doi.org/10.1186/ISRCTN19682964)

## 1.0 Introduction

Diabetes mellitus is a highly prevalent disorder and a leading cause of morbidity and mortality (X. Lin et al., 2020). In 2022 an estimated 828 million individuals worldwide were living with diabetes, with type 2 diabetes (T2D) accounting for over 80% of cases (Ong et al., 2023; Zhou et al., 2024). This figure is expected to increase to 1.3 billion by 2050, with rates rising higher in children and those under 40 years of age (Ahmad et al., 2022; Lascar et al., 2018; Ong et al., 2023). T2D is associated with a myriad of long-term health conditions, including vascular disease, heart disease, kidney disease, retinopathy, and neuropathy (Ahmad et al., 2022). Within the UK, T2D poses a substantial financial burden on the National Health Service (NHS), consuming around 10% of the annual healthcare budget (Whicher et al., 2020).

Sleep plays a key role in cardiometabolic function. It is well established from prospective data that sleep duration (both short and long) is associated with a greater risk of diabetes (Cappuccio et al., 2010; Chao et al., 2011; Shan et al., 2015). Further, experiencing insufficient sleep due to later sleep onset times and an evening chronotype is associated with increased risk of T2D (Ma et al., 2022; Shen et al., 2025). Evidence points to a chronic pattern of sleep debt and circadian rhythm disturbance, also known as social jetlag, to which evening chronotypes are predisposed, conferring a greater risk of T2D and non-diabetic hyperglycaemia in younger adults and poorer glycaemic control (higher HbA_1c_) in adults with T2D (Kelly et al., 2020; Koopman et al., 2017). In non-obese, healthy participants, experimental sleep restriction and sleep fragmentation drive change in factors associated with diabetic status, including increased hunger, reduced insulin sensitivity / increased insulin resistance, increased postprandial glucose surges, and increased cortisol (Reynolds et al., 2012; Stamatakis & Punjabi, 2010; Sweeney et al., 2017; Zhu et al., 2019; Zuraikat et al., 2024). Conversely, experimental sleep extension in short sleepers improves metabolic parameters and weight reduction (Leproult et al., 2015; So-Ngern et al., 2019; Tasali et al., 2022).

Insomnia is a prominent comorbidity in those with T2D, affecting 39% of patients (Schipper et al., 2021; Koopman et al. 2020). Conversely, insomnia increases risk for developing T2D (Wright et al., 2025). This is risk is heightened in younger participants (under 40 years of age);(Lin et al., 2018). Mendelian randomisation also shows that genetic instruments for frequent insomnia symptoms are causally related to elevated glycated haemoglobin (HbA_1C_) (Liu et al., 2022; Ma et al., 2022; Yuan & Larsson, 2020) and that insulin resistance is a significant mediator of the relationship between insomnia and T2D(Wang et al., 2023).

If insomnia is causally related to hyperglycaemia, then effective intervention could improve metabolic health. Few studies have evaluated CBT in individuals with insomnia and T2D (Mostafa et al., 2025). Findings are mixed across studies but generally show improvement in HbA_1C_ and C-reactive protein (CRP), albeit with small effect sizes (Mostafa et al., 2025; Savin et al., 2023). Small and heterogeneous samples and poor treatment engagement were all identified as factors influencing the quality and robustness of evidence to date. To our knowledge, no study has assessed the effects of sleep intervention in non-diabetic hyperglycaemia (NDH), a key risk state for diabetes characterised by raised glycated haemoglobin (HbA1c) levels below the diagnostic threshold for.

To address this gap, we conducted a proof-of-principle feasibility trial of digitial cognitive behavioural therapy (dCBT) in individuals with insomnia and NDH to inform a future large-scale RCT. The primary aim of the study was to evaluate the feasibility of recruiting eligible participants with both insomnia and NDH from primary care to the trial. Secondary outcomes were to explore adherence to the intervention protocol and provide proof-of-principle data regarding changes in sleep and metabolic outcomes.

## 2.0 Materials and Methods

## 2.1 Participants

Ten primary care practices within Oxfordshire, UK, were identified and approached to identify participants. Practice medical records were searched using a preset screening algorithm to identify potential participants. Potentially eligible participants were identified as being aged 18 years or over, having either an HbA1c of 6-6.4% within the past 12 months or a diagnosis of NDH (pre-diabetes), no current diagnosis of diabetes or current use of diabetic medication, no recorded diagnosis of sleep apnoea, and no current hypnotic prescription.

All patients within the practice who met these initial search criteria were sent a letter and participant information sheet and invited to complete an online screening questionnaire (Qualtrics, Provo, UT) to further assess eligibility. Inclusion criteria included meeting DSM-5 criteria for insomnia disorder according to the Sleep Condition Indicator (SCI (Espie et al., 2014)), having reliable internet access, and being able to understand the study instructions in English. Participants were excluded if they had evidence of another sleep disorder, including obstructive sleep apnea (OSA, assessed with the STOP-BANG (Chung et al., 2016)), narcolepsy, restless legs syndrome (RLS)/periodic limb movement disorder, circadian rhythm sleep-wake disorder, or parasomnia (Espie, 2024; Wilson et al., 2010); psychosis or epilepsy; prescribed diabetes or sleep medication; engaged in shift work; dementia or mild cognitive impairment; suicidal ideation with intent; pregnant or planning pregnancy; active cancer; planned surgery in the next 2 months; life expectancy of less than 1 year; previous or current access to the digital CBTi programme (Sleepio™); currently receiving psychological therapy for insomnia or engaged in another sleep intervention trial; or had an allergy to hypoallergenic adhesive plasters.

The study was approved by the Health Research Authority (IRAS 266313) and the allocated Research Ethics Committee (East of England – Essex Research Ethics Committee, 20/EE/0046) and was prospectively registered on the ISRCTN (ISRCTN19682964)

### 2.2 Procedures

All study procedures took place at the NIHR Oxford University Hospitals Biomedical Research Centre Clinical Research Unit, within the Oxford Centre for Diabetes, Endocrinology, and Metabolism, UK (OCDEM CRU). Once eligibility was confirmed, participants were invited to a baseline research appointment with a research nurse where they gave informed consent to the trial, had a blood sample drawn, completed a cognitive task and questionnaires, were fitted with a continuous glucose monitor (CGM: Dexcom, California, USA), were given a sleep diary (Consensus Sleep Diary: (Carney et al., 2012)), and an a wrist-worn actigraph “actiwatch” (MotionWatch8, CamNtech, UK). A research nurse completed CGM placement to ensure standardisation between participants, affixing the device to the participant’s abdomen.

For the next 7 days, participants wore the CGM and actiwatch and completed the sleep diary. They then returned to the research unit to have the CGM removed and return the actiwatch and sleep diary. Immediately following this, participants were given access to the sleep intervention to complete over 10 weeks. At Week 11, participants returned to OCDEM CRU for the post-assessment visit, which followed the same procedures as the baseline visit. Participants were reimbursed with a £50 voucher for participation in the trial, alongside travel expenses. For the study flow, see Figure 1.

**Figure 1:**
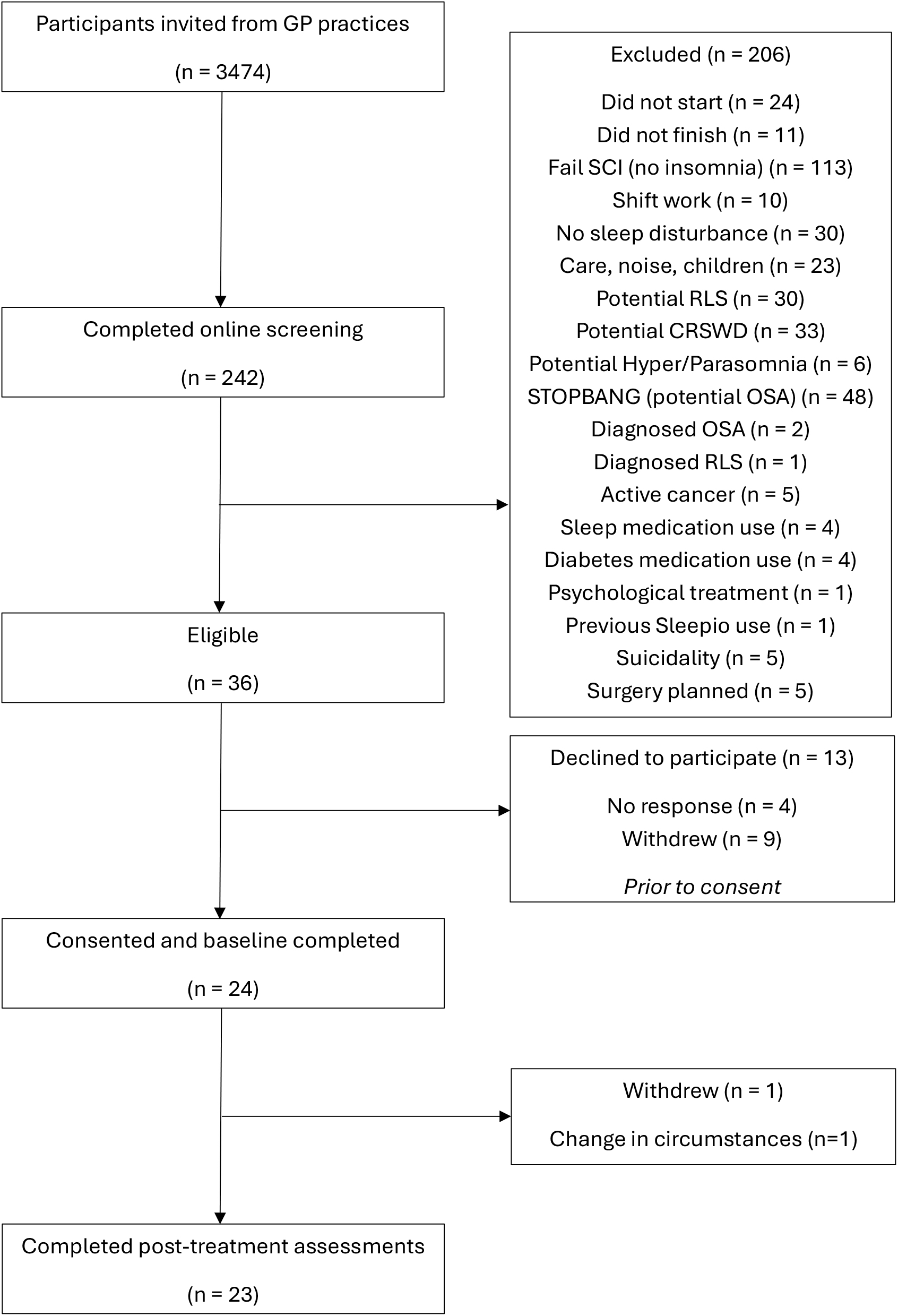
STUDY FLOW. Diagram demonstrating the flow of procedures and measurements for the SleepTMH study

### 2.3 Intervention

The digital cognitive behavioural therapy for insomnia (dCBTIi) intervention was delivered using the Sleepio™ programme, a NICE-approved and FDA-cleared fully automated programme, shown to be clinically effective (Espie et al., 2012). The six-session programme was completed independently over a ten-week period. The treatment includes a behavioural component (sleep restriction, stimulus control, and relaxation), a cognitive component (paradoxical intention, cognitive restructuring, mindfulness, positive imagery, and putting the day to rest) and an educational component (psycho-education and sleep hygiene). Each session lasts for around 20 minutes and the content of the programme is presented by an animated virtual therapist. The programme includes a daily sleep diary which is used by the programme to create personalised help and prescribed sleep-wake schedules. The system provided online analytics, which can be used to monitor engagement by assessing how many sessions were completed and the number of weeks to complete the course.

### 2.4 Outcome Measures

#### 2.4.1. Primary Outcome - Feasibility

The primary outcome was the feasibility of recruiting eligible participants from a primary care setting. For this, recruitment rates per month were calculated.

#### 2.4.2. Secondary Outcomes

##### 2.4.2.1. Acceptability

Acceptability was indexed by the proportion of participants who completed at least four of the six intervention sessions within the ten-week period.

##### 2.4.2.2. Treatment Satisfaction

At the week 11 visit, participants completed a treatment satisfaction questionnaire consisting of two questions assessed on a 5-point Likert scale. Participants were asked “Overall, how satisfied are you with the sleep treatment you received?” scored from 0 ‘Very Dissatisfied’ to 4 ‘Very satisfied’. Perceived sleep improvement was captured by the item “To what extent did the sleep treatment improve your sleep?” scored from 0 ‘Not at all’ to 4 ‘A lot’.

##### 2.4.2.3. Proof of Principle Outcome Measures

Proof-of-principle outcome data were collected to inform a full RCT. Measures were collected at week 0-1 (baseline) and weeks 11-12 (post-assessments).

Insomnia severity was measured by the Insomnia Severity Index (ISI: (Morin, 1993)), depressive symptomology was measured by the Centre for Epidemiologic Studies Depression Scale (CES-D: (Radloff, 1977)), chronotype was measured by the Morningness-Eveningness Questionnaire (reduced version, rMEQ: (Adas & Alwrall, 1991)), health-related quality of life was measured by EuroQol (EQ-5D-3L: (Kind et al., 1998; The EuroQoL Group, 1990)), and overall self-rated health was assessed by the visual analogue scale from the EQ-5D (EQ-5D-3L VAS).

Subjective sleep was assessed for 7 days at pre- and post-treatment using the Consensus Sleep Diary (CSD: (Carney et al., 2012)(Carney et al., 2012)). Outcomes were: time in bed (TIB); total sleep time (TST); wake after sleep onset (WASO); sleep efficiency (SE); sleep onset latency (SOL); and sleep quality (scored on a 5-point Likert scale from 0 ‘very poor’ to 4 ‘very good’). We also assessed cognitive arousal each morning using item five (“last night, as you were attempting to fall asleep or return to sleep was your mind mentally alert and active”) from the pre-sleep arousal scale (PSAS: (Nicassio et al., 1985)(Nicassio et al., 1985)) scored on a 5-point Likert scale from 0 ‘not at all’ to 4 ‘extremely’.

Estimated objective sleep parameters and rest-activity rhythms were derived from actigraphy (MotionWatch 8, CamNtech) recorded over a seven-day collection period at 30-second epochs on a medium sensitivity. Outcomes were: TIB; TST; WASO; SE; SOL; relative amplitude of the rest activity cycle (RA); interdaily stability of the rest activity cycle (IS); and intradaily variability of the rest activity cycle (IV).

A fasted blood sample was collected from participants at week 0 and week 11 assessments. Participants were instructed to fast from 10pm the night before their visit. Each blood sample was 9.5ml of whole blood. Blood samples were analysed for HbA_1c_, insulin, glucose, non-esterified fatty acids (NEFA), glycerol, lactate, urea, triglycerides, low-density lipoprotein cholesterol, high-density lipoprotein cholesterol, beta-hydroxybutyrate, apolipoprotein-B, alanine transaminase, and C-reactive protein.

Free-living interstitial glucose concentrations were recorded with a continuous glucose monitoring system (CGM; Dexcom G6). The CGM was affixed to the participant’s abdomen and measured interstitial glucose levels every 5 minutes over a one-week recording period, concurrent with the measurements above. The CGM device was utilised in a blinded mode, meaning participants did not receive live feedback on their glucose levels. However, for safety, alarms were set to sound if blood sugar levels reached an extreme ‘high’ or ‘low’ during the recording period.

Working memory performance was assessed at week 0 and week 11 using the visuospatial working memory task (VSTM) (Zhao et al., 2025)(Zhao et al., 2025). The VSTM takes around 10 minutes to complete. Participants are presented with 1 or 3 fractal patterns on the screen. After a 4-second delay, one of the previously presented fractal patterns are displayed alongside a foil fractal (one not presented previously). Participants are required to identify the true presented fractal pattern and then move the fractal to the position where it originally appeared on the screen. This task probes both identification memory and localisation memory (number correctly identified fractals, identification reaction time, and localisation reaction time).

##### 2.4.2.4. Adverse events

Participants were asked to report on accidents and injuries occurring within the past two months, both pre- and post-intervention. This 5-item questionnaire included 3 binary (yes/no) items, seeking responses to whether the participant had experienced a work-related accident, a motor vehicle accident, or a near-miss driving incident within the past 2 months. The final two items asked the participant to report how many times they had fallen asleep while driving and how many times they had experienced a fall within the past two months.

During study visits, nurses enquired about any serious adverse events, defined as any untoward medical occurrence that results in death, is life-threatening, requires inpatient hospitalisation or prolongation of existing hospitalisation, results in persistent or significant disability/incapacity, consists of a congenital anomaly or birth defect.

### 3.0 Statistical analysis

A statistical analysis plan was drafted and finalised prior to final data lock. Data were analysed by a statistician (VH) using STATA (StataCorp. 2025. Stata Statistical Software: Release 19. College Station, TX: StataCorp LLC). Initial processing of actigraphy data was conducted in the proprietary MotionWare software. CGM data was processed using the iglu R package (Broll et al., 2021)(Broll et al., 2021) and consistent with the recommendations of the Lancet Diabetes & Endocrinology consensus statement (Battelino et al., 2023)(Battelino et al., 2023).

The primary outcome measure included all participants who consented to the trial. Participants who withdrew from the trial were included in the analysis of outcomes up to the point of withdrawal. Participants were included in the analysis regardless of whether they engaged with the dCBTi intervention.

For the primary outcome, total number of participants and total recruitment period were used to calculate the average recruitment rate per month. Intervention engagement was calculated as the proportion of participants who completed four or more dCBTi sessions. Treatment satisfaction information was grouped into three levels: dissatisfied, neutral, satisfied.

Continuous outcomes, comprising diary, actigraphy, questionnaire, glucose and other blood biochemical variables, were evaluated for average change from pre-to-post treatment. Consistent with the aims of a feasibility trial, we reported 95% CI for change but did not perform formal statistical significance testing. Cohen’s d effect size was calculated using the within-subject treatment effect and the standard deviation of change score.

## 4.0 Results

### 4.1 Feasibility

Ten local primary care practices identified 3474 participants as potentially meeting eligibility criteria and were sent a participant information sheet. Of this number, 242 (7%) completed the online screening and 36 were considered eligible for the study. Nine participants withdrew prior to consent, and four did not respond to further contact, leaving 24 participants enrolled. All 24 participants commenced the intervention and completed baseline procedures. One participant withdrew due to changes in personal circumstances and declined to complete post-assessment outcome measures, resulting in 23 participants (96%) completing the study (see Figure 2 for the Consolidated Standards of Reporting Trials (CONSORT) flow diagram). Study participants included 17 women (70.8%), mean age 65.5 years (SD = 12.4), CES-D mean of 12.8 (SD = 9.6), ISI score of 15.2 (SD = 4.9), insomnia duration of 15 years (SD = 16.78), and a mean fasting glucose level of 4.9 mmol/l (SD = 1.1) (Table 1).

**Table 1:**
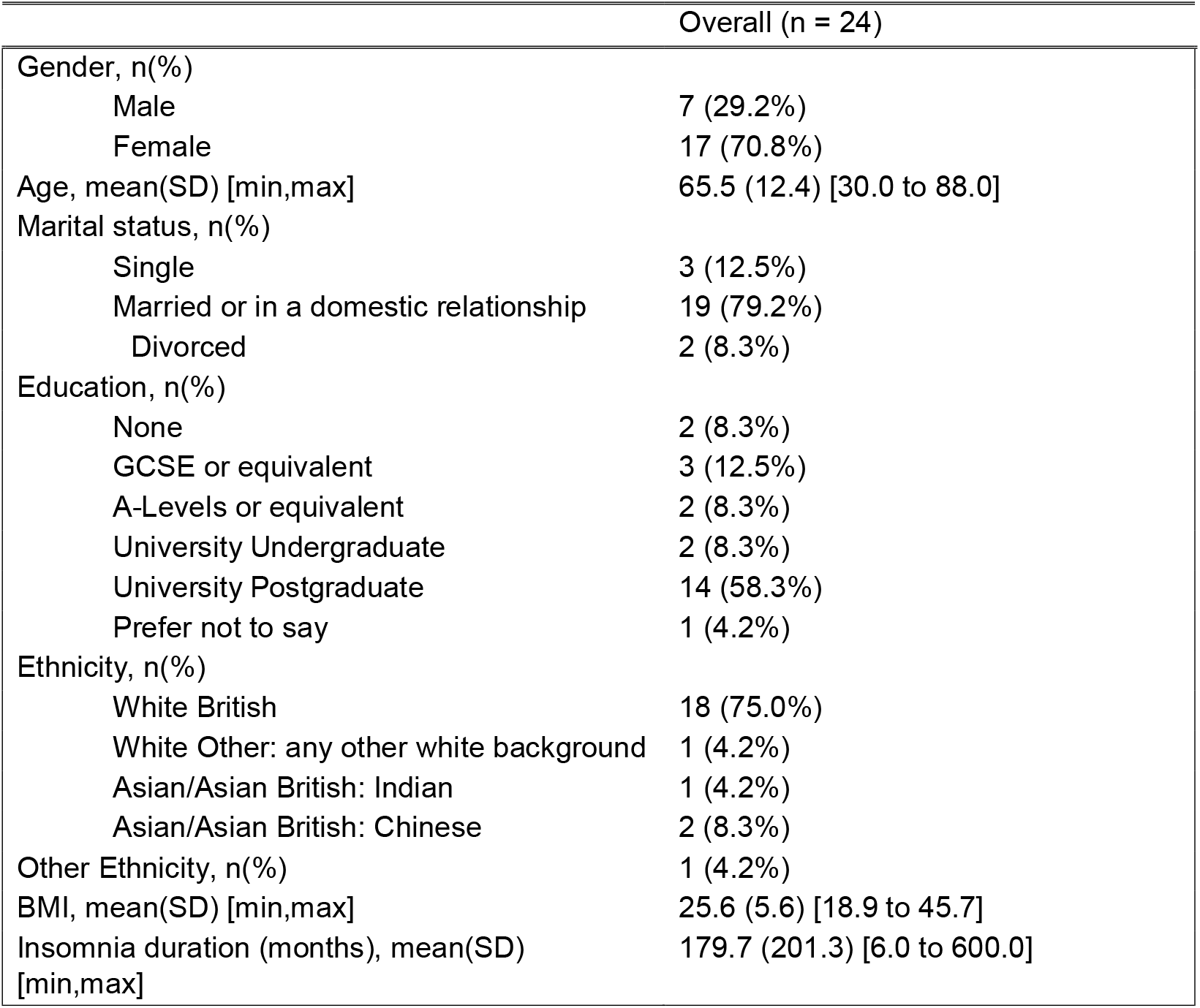
Baseline characteristics of the participants.

**Figure 2:**
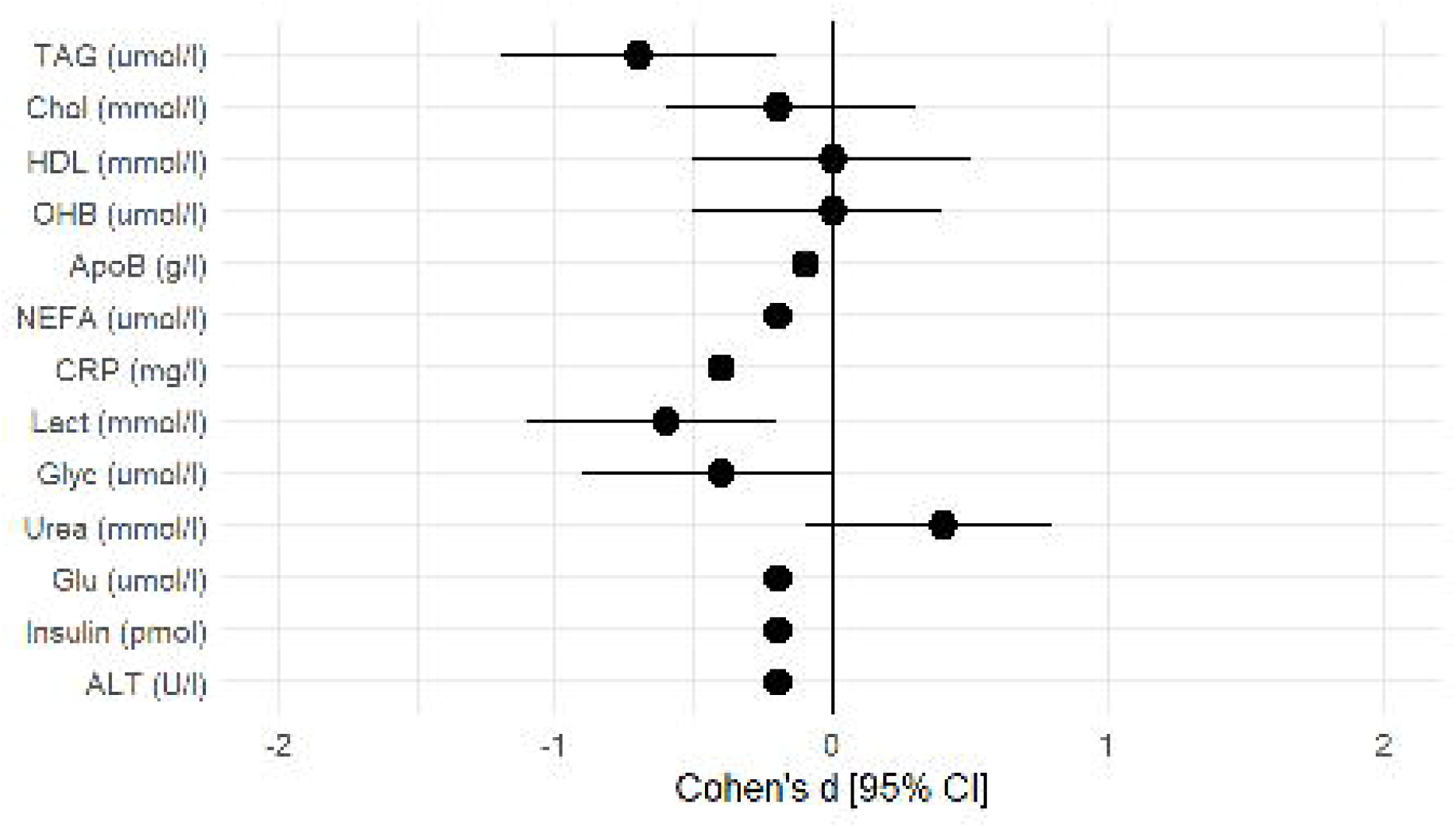
CONSORT DIAGRAM. Consolidated Standards of Reporting Trials (CONSORT) flow diagram. Participants could be excluded due to multiple reasons and therefore the number of participants excluded is less than the summation of all the reasons for exclusion

Recruitment took place over 16 months, with an average of 1.5 participants recruited per month. The majority of recruitment occurred in the final 4 months of the study, with an average of 5 participants recruited per month (Supplemental Figure 1). Slow initial recruitment was due, in part, to a staggered resumption of activities following the COVID-19 pandemic.

### 4.2 Acceptability

All 24 participants started session 1 of the intervention. Twenty (83.3%) participants completed at least 4 intervention sessions, with two participants completed five sessions and 18 (75%) participants attending all six sessions. Overall, the mean number of sessions attended was 5.3 (SD = 1.5). Seventeen participants (70.8%) were satisfied with the treatment, 6 (25%) were neutral, and no participant reported feeling dissatisfied. Eight participants (33.3%) reported ‘quite a bit’ or ‘a lot’ of improvement in their sleep following treatment. 7 (29.2%) reported ‘somewhat’ of an improvement, and 8 (33.3%) reported ‘not at all’ or ‘very little’ improvement.

### 4.3 Proof of principle outcome measures

Improvements in questionnaire-based outcomes were apparent in some measures from pre-to post-treatment (Table 2), including medium-to-large reductions in insomnia symptoms (ISI) [−4.7(95%CI:−6.2 to −3.2), *d*=−1.4] and depressive symptoms (CES-D) [−2.7(95%CI:−5.1 to −0.2), *d*=−0.5]. There was no evidence of change in chronotype (rMEQ) or health-related quality of life (EQ5D-3L).

**Table 2:**
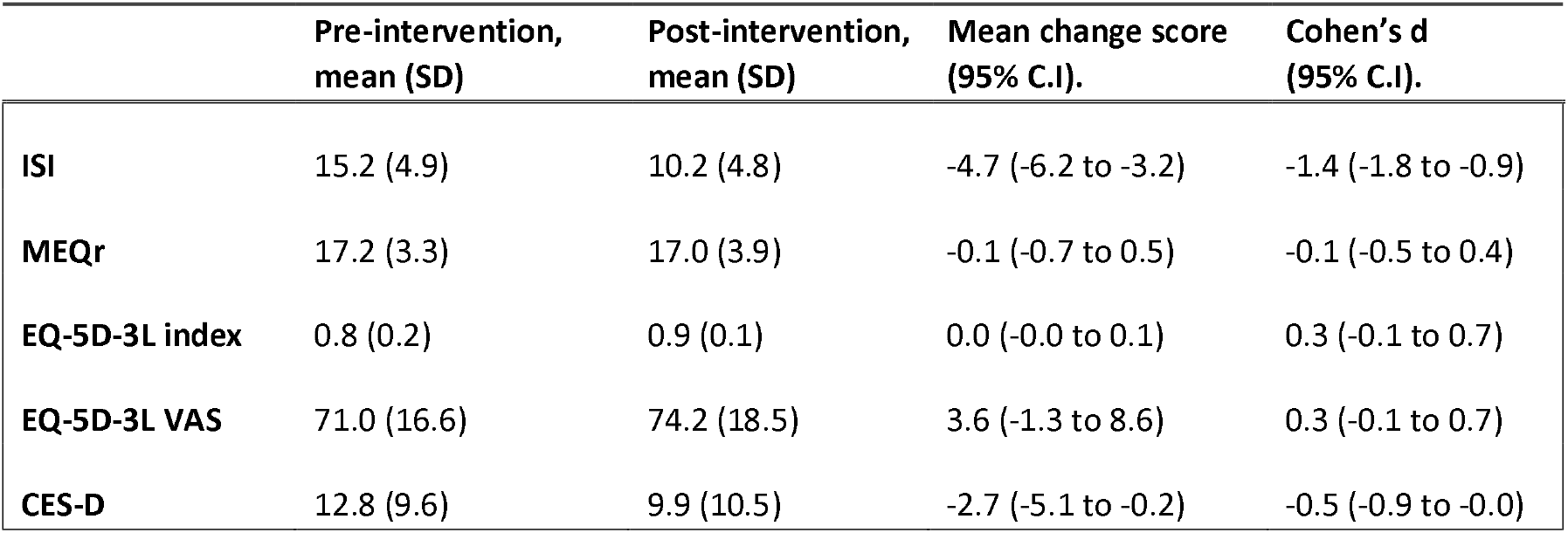
Pre-intervention and post-intervention means, standard deviations (SD), pre-to-post change scores, and effect sizes (Cohen’s d) for the questionnaire items. 95% confidence intervals (C.I) are presented for the mean change and the effect size. ISI: Insomnia Severity Index. MEQr: Morningness-Eveningness Questionnaire reduced form.EQ-5D-3L: EuroQol 5 Dimension. CES-D: Centre for Epidemiologic Studies Depression Scale.

There tended to be greater sleep improvement in diary outcomes compared to the actigraphy data (Tables 3 and 4 and Supplemental Figure 2). For TIB, the effect was consistent across both actigraphic and diary measures, with a medium reduction of 33.2 minutes (*d* = −0.5). For total sleep time (TST), actigraphy data revealed a decrease of 18.5 minutes (*d* = −0.5), whereas the sleep diary data showed an increase of 21.1 minutes (*d* = 0.5). Self-reported SOL (*d* = −0.9), SE (*d* = 0.8), sleep quality (*d* = 0.6) and cognitive arousal (*d* = −0.6) improved from pre-to-post intervention. There were no other changes in actigraphy-defined rest-activity or sleep continuity outcomes.

**Table 3:** Pre-intervention and post-intervention means, standard deviations (SD), pre-to-post change scores, and effect sizes (Cohen’s d) for the actigraphy outcome measures, averaged over the week. 95% confidence intervals (C.I) are presented for the mean change and the effect size. TIB: Time In Bed (decimal minutes). TST: Total Sleep Time (decimal minutes). WASO: Wake After Sleep Onset (decimal minutes). SE: Sleep Efficiency (%). SOL: Sleep Onset Latency (decimal minutes). RA: Relative Amplitude of the rest activity cycle (non-parametric circadian rhythm analysis). IS: Interdaily Stability of the rest activity cycle (non-parametric circadian rhythm analysis). IV: Intradaily Variability of the rest activity cycle (non-parametric circadian rhythm analysis).

|  | Pre-intervention,<br>mean (SD) | Post-intervention,<br>mean (SD) | Mean change score<br>(95% C.I.) | Cohen's d<br>(95% C.I.) |
| --- | --- | --- | --- | --- |
| **TIB** | 540.5 (64.3) | 508.3 (46.3) | -33.2 (-61.9 to -4.4) | -0.5 (-1.0 to -0.1) |
| **TST** | 444.2 (45.3) | 424.9 (43.3) | -18.5 (-36.5 to -0.5) | -0.5 (-0.9 to -0.0) |
| **WASO** | 72.1 (35.2) | 62.4 (25.0) | -8.7 (-22.5 to 5.1) | -0.3 (-0.7 to 0.2) |
| **SE** | 82.7 (7.6) | 83.8 (6.0) | 1.3 (-0.5 to 3.2) | 0.3 (-0.1 to 0.8) |
| **SOL** | 9.4 (12.4) | 9.6 (8.6) | 0.3 (-2.6 to 3.2) | 0.0 (-0.4 to 0.5) |
| **RA** | 0.868 (0.086) | 0.885 (0.067) | 0.017 (-0.029 to 0.063) | 0.3 (-0.2 to 0.7) |
| **IS** | 0.544 (0.123) | 0.535 (0.105) | -0.009 (-0.077 to 0.060) | -0.2 (-0.7 to 0.2) |
| **IV** | 0.907 (0.210) | 0.942 (0.149) | 0.035 (-0.074 to 0.144) | 0.2 (-0.3 to 0.6) |

**Table 4:**
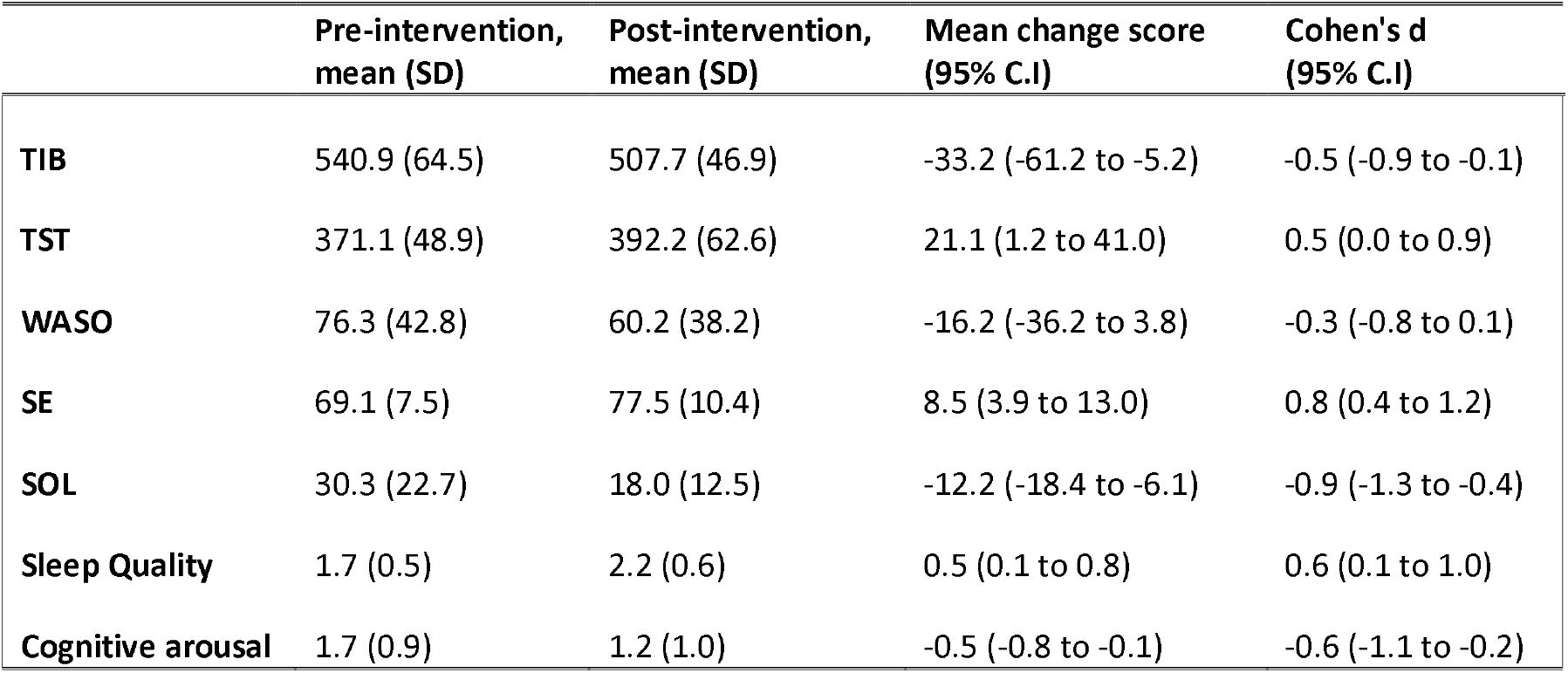
Pre-intervention and post-intervention means, standard deviations (SD), pre-to-post change scores, and effect sizes (Cohen’s d) for the sleep diary outcome measures, averaged over the week. 95% confidence intervals (C.I) are presented for the mean change and the effect size. TIB: Time In Bed (decimal minutes). TST: Total Sleep Time (decimal minutes). WASO: Wake After Sleep Onset (decimal minutes). SE: Sleep Efficiency (%). SOL: Sleep Onset Latency (decimal minutes). Sleep Quality: scored on a 5-point Likert scale from 0 ‘very poor’ to 4 ‘very good’, with higher average scores indicating better self-reported sleep quality. Cognitive arousal: scored on a 5-point Likert scale from 0 ‘not at all’ to 4 ‘extremely’, with higher scores indicating greater pre-sleep cognitive arousal.

There was a trend towards improvement in most of the fasting blood outcome measures (Table 5 and Figure 3). There was a medium-to-large reduction in triglyceride (TAG) levels, as well as medium reductions in lactate (Lact) and glycerol (Glyc) levels. A laboratory processing error resulted in the loss of HbA_1c_ data.

**Table 5:** Pre-intervention and post-intervention means, standard deviations (SD), pre-to-post change scores, and effect sizes (Cohen’s d) for the fasting blood sample outcome measures. 95% confidence intervals (C.I) are presented for the mean change and the effect size. TAG: Triglycerides. Chol: Cholesterol. HDL: High Density Lipoprotein. OHB: β-hydroxybutyrate. ApoB: Apolipoprotein B. NEFA: Non-Esterified Fatty Acids. CRP: C-Reactive Protein. Lact: Lactate. Glyc: Glycerol. Glu: Glucose. ALT: Alanine Aminotransferase

|  | Pre-intervention,<br>mean (SD) | Post-intervention,<br>mean (SD) | Mean change score<br>(95% C.I.) | Cohen's d<br>(95% C.I.) |
| --- | --- | --- | --- | --- |
| **TAG (umol/l)** | 930.0 (366.3) | 780.0 (253.5) | -148.7 (-245.9 to -51.4) | -0.7 (-1.2 to -0.2) |
| **Chol (mmol/l)** | 4.4 (1.2) | 4.2 (1.0) | -0.2 (-0.8 to 0.4) | -0.2 (-0.6 to 0.3) |
| **HDL (mmol/l)** | 2.6 (1.1) | 2.6 (1.2) | -0.0 (-0.5 to 0.5) | -0.0 (-0.5 to 0.5) |
| **OHB (umol/l)** | 155.3 (199.0) | 146.1 (144.0) | -3.3 (-121.2 to 114.6) | -0.0 (-0.5 to 0.4) |
| **ApoB (g/l)** | 0.8 (0.2) | 0.7 (0.2) | -0.0 (-0.1 to 0.1) | -0.1 (-0.6 to 0.3) |
| **NEFA (umol/l)** | 664.2 (416.9) | 567.5 (253.3) | -87.4 (-263.6 to 88.8) | -0.2 (-0.7 to 0.2) |
| **CRP (mg/l)** | 4.2 (2.8) | 3.2 (0.5) | -1.0 (-2.2 to 0.2) | -0.4 (-0.8 to 0.1) |
| **Lact (mmol/l)** | 1.2 (0.5) | 0.9 (0.3) | -0.3 (-0.5 to -0.1) | -0.6 (-1.1 to -0.2) |
| **Glyc (umol/l)** | 63.0 (35.2) | 51.3 (25.6) | -13.1 (-26.7 to 0.5) | -0.4 (-0.9 to 0.0) |
| **Urea (mmol/l)** | 5.8 (1.8) | 6.1 (1.6) | 0.5 (-0.1 to 1.1) | 0.4 (-0.1 to 0.8) |
| **Glu (mmol/l)** | 4.9 (1.1) | 4.7 (0.7) | -0.2 (-0.7 to 0.3) | -0.2 (-0.6 to 0.3) |
| **Insulin (pmol)** | 27.9 (23.3) | 31.6 (31.4) | -3.3 (-10.1 to 3.5) | -0.2 (-0.7 to 0.2) |
| **ALT (U/l)** | 46.2 (26.9) | 45.0 (17.5) | -3.9 (-14.0 to 6.3) | -0.2 (-0.6 to 0.3) |

**Figure 3:**
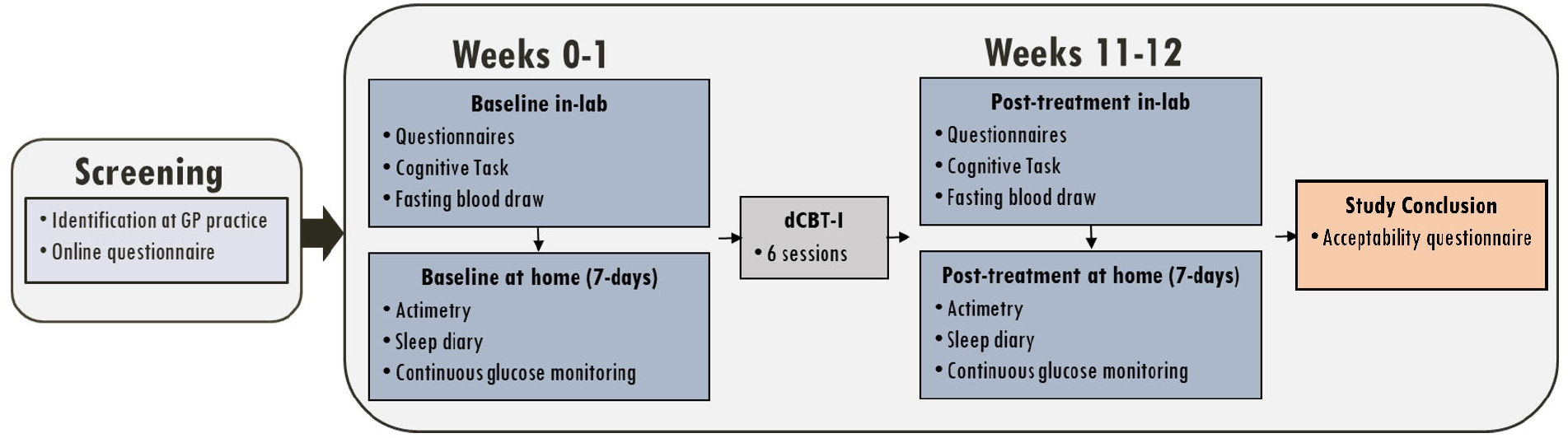
FASTING BLOOD FOREST PLOT. Forest plot showing the 95% confidence intervals of the effect sizes for the fasting blood sample outcome measures. TAG: Triglycerides. Chol: Cholesterol. HDL: High Density Lipoprotein. OHB: β-hydroxybutyrate. ApoB: Apolipoprotein B. NEFA: Non-Esterified Fatty Acids. CRP: C-Reactive Protein. Lact: Lactate. Glyc: Glycerol. Glu: Glucose. ALT: Alanine Aminotransferase

There were no clear pre-to-post differences in visuospatial working memory (VSTM) or continuous glucose measures (Supplemental Tables 1 and 2).

### 4.4 Adverse events

No serious adverse events were reported to the trial team. There was no evidence of increased rates of pre-defined adverse events (work-related accidents, motor vehicle accidents, near-misses while driving, falling asleep while driving, or falls) following the intervention period (Supplemental Table 3).

## 5.0 Discussion

This study provides preliminary data to support a future trial of dCBTi in individuals with NDH. Regarding the primary outcome, recruitment rate was initially slower than expected, which we attribute in part to the COVID-19 pandemic and challenges in resuming clinical research activities. Initial recruitment saw three participants enrolled between August 2022 and March 2023 from four primary practices. The addition of six practices from August 2023 to November 2023 saw 21 participants enrolled at a rate of 5 participants per month. Treatment engagement was excellent (83.3% completed ≥4/6 sessions), treatment satisfaction was high (n=17 satisfied, n=6 neutral), and follow-up rates were high (96%).

Consistent with the broader literature regarding digital sleep interventions, mean change, confidence intervals, and effect size estimates indicated positive improvements in sleep and depression symptomology (Espie et al., 2012; Henry et al., 2021; Kyle et al., 2020; Tamm et al., 2025)(Espie et al., 2012; Henry et al., 2021; Kyle et al., 2020; Tamm et al., 2025). Subjective sleep parameters showed greater improvement than actigraphy-defined sleep. Although no inferential statistics were conducted, we observed a potential reduction in markers of lipolysis, suggesting improved adipose tissue function and a trend towards reduced systemic inflammation (CRP) following dCBTi. A larger, adequately powered, randomised trial is now indicated to confidently appraise the impact of insomnia treatment on key metabolic outcomes in those at risk of developing T2D.

Several factors should be considered when interpreting our findings. The primary focus of this single-centre, non-randomised study was on feasibility and acceptability; therefore, no conclusions can be drawn regarding efficacy. Our recruited sample was highly educated and had a relatively low BMI (25.6) compared with the broader NDH population (e.g., mean of 31.1 within the UK national diabetes prevention programme (Ross et al., 2022)(Ross et al., 2022). Our decision to exclude people with a diagnosis of sleep apnoea or those screening positive for OSA (on the STOP-BANG) likely contributed to sample characteristics. Eligibility screening for this study was conducted entirely via questionnaires and, therefore, may have reduced diversity in the sample due to exclusion through over-sensitivity and low specificity of the measures. Mean baseline fasting glucose levels turned out to be in the normal range at 4.9mmol/L (NDH range is 5.6-6.9mmol/L), potentially limiting scope for improvement post-intervention. This may relate to our inclusion criteria of having an HbA_1c_ level of 6-6.4% within the past 12 months or a diagnosis of pre-diabetes. Future studies may wish to employ higher specificity screening measures, including home-based assessments of respiratory sleep disorders, and conduct a screening measure of blood sugar levels to ensure NDH status prior to study intake. Participation in other lifestyle interventions was not an exclusion; therefore, participants may have been simultaneously engaged in other interventions whilst in this trial, which was not monitored or recorded.

Several countries have introduced lifestyle intervention programmes aimed at preventing the conversion from NDH to T2D (for examples see UK (Ross et al., 2022)(Ross et al., 2022) and USA (The Diabetes Prevention Program (DPP) Research Group, 2002)(The Diabetes Prevention Program (DPP) Research Group, 2002)). These programmes primarily focus on improving glycaemic outcomes through caloric control and increasing physical activity. Emerging data identify sleep as a potentially important modifiable factor with causal relevance to metabolic health (Che et al., 2021; Liu et al., 2022)(Che et al., 2021; Liu et al., 2022). This feasibility study has demonstrated that a digital sleep intervention for individuals with NDH is both feasible and acceptable. Early signals on sleep, inflammatory, and cardiometabolic outcomes now require evaluation in large-scale randomised controlled trials.

## Data Availability

All data produced in the present study are available upon reasonable request to the authors

## Acknowledgements

This study was funded by the National Institute for Health and Care Research (NIHR) Oxford Health Biomedical Research Centre (ref: NIHR203316) and the NIHR Oxford Biomedical Research Centre (ref: BRC-1215-20008, Cluster Award 2019-2020). AF is supported by the NIHR Oxford BRC. RS, DR, PG, CE and SDK are supported by the NIHR Oxford Health BRC. The views expressed are those of the author(s) and not necessarily those of the NIHR or the Department of Health and Social Care.

## Author Contribution

**RS**: conceptualisation (equal); data curation (lead); investigation (lead); methodology (equal); project administration (lead); original draft (lead); writing – review and editing (equal). **DR**: funding acquisition (equal); conceptualisation (equal); writing – review and editing (equal). **AF**: funding acquisition (equal); conceptualisation (equal); writing – review and editing (equal). **PCEG**: writing – conceptualisation (supporting); visualisation (lead). **VH**: formal analysis (lead); writing – review and editing (equal). **FK**: funding acquisition (equal); conceptualisation (equal); writing – review and editing (equal). **DM**: investigation (support). **TM**: investigation (support). **NMM:** data curation (support); investigation (support); project administration (support); writing – review and editing (equal). **JWT**: funding acquisition (equal); conceptualisation (equal); writing – review and editing (equal). **CE**: investigation (support); writing – review and editing (equal). **SDK**: funding acquisition (lead); conceptualisation (equal); supervision (lead); writing – review and editing (equal).

## References

Adas, A., & Alwrall, H. (1991). HORNE & OSTBERG MORNINGNESS-EVENINGNESS QUESTIONNAIRE: A REDUCED SCALE. Personality and Individual Differences, 12(3), 241–253.

Ahmad, E., Lim, S., Lamptey, R., Webb, D. R., & Davies, M. J. (2022). Type 2 diabetes. In The Lancet (Vol. 400, Issue 10365, pp. 1803–1820). Elsevier B.V. 10.1016/S0140-6736(22)01655-5

Battelino, T., Alexander, C. M., Amiel, S. A., Arreaza-Rubin, G., Beck, R. W., Bergenstal, R. M., Buckingham, B. A., Carroll, J., Ceriello, A., Chow, E., Choudhary, P., Close, K., Danne, T., Dutta, S., Gabbay, R., Garg, S., Heverly, J., Hirsch, I. B., Kader, T., … Phillip, M. (2023). Continuous glucose monitoring and metrics for clinical trials: an international consensus statement. The Lancet Diabetes & Endocrinology, 11(1), 42–57. 10.1016/S2213-8587(22)00319-9

Broll, S., Urbanek, J., Buchanan, D., Chun, E., Muschelli, J., Punjabi, N. M., & Gaynanova, I. (2021). Interpreting blood GLUcose data with R package iglu. PLoS ONE, 16(4 April). 10.1371/journal.pone.0248560

Cappuccio, F. P., Lanfranco D’elia,;, Strazzullo,; Pasquale, & Miller, M. A. (2010). Sleep Duration and All-Cause Mortality: A Systematic Review and Meta-Analysis of Prospective Studies. In SLEEP (Vol. 33, Issue 5). https://academic.oup.com/sleep/article/33/5/585/2454478

Carney, C. E., Buysse, D. J., Ancoli-Israel, S., Edinger, J. D., Krystal, A. D., Lichstein, K. L., & Morin, C. M. (2012). The consensus sleep diary: Standardizing prospective sleep self-monitoring. In Sleep (Vol. 35, Issue 2, pp. 287–302). 10.5665/sleep.1642

Chao, C. Y., Wu, J. S., Yang, Y. C., Shih, C. C., Wang, R. H., Lu, F. H., & Chang, C. J. (2011). Sleep duration is a potential risk factor for newly diagnosed type 2 diabetes mellitus. Metabolism: Clinical and Experimental, 60(6), 799–804. 10.1016/j.metabol.2010.07.031

Che, T., Yan, C., Tian, D., Zhang, X., Liu, X., & Wu, Z. (2021). The Association Between Sleep and Metabolic Syndrome: A Systematic Review and Meta-Analysis. Frontiers in Endocrinology, 12. 10.3389/fendo.2021.773646

Chung, F., Abdullah, H. R., & Liao, P. (2016). STOP-bang questionnaire a practical approach to screen for obstructive sleep apnea. Chest, 149(3), 631–638. 10.1378/chest.15-0903

Espie, C. A. (2024). The Clinician’s Guide to Cognitive and Behavioural Therapeutics (CBTx) for Insomnia. Cambridge University Press. 10.1017/9781108989480

Espie, C. A., Kyle, S. D., Hames, P., Gardani, M., Fleming, L., & Cape, J. (2014). The Sleep Condition Indicator: A clinical screening tool to evaluate insomnia disorder. BMJ Open, 4(3), 1–6. 10.1136/bmjopen-2013-004183

Espie, C. A., Kyle, S. D., Williams, C., Ong, J. C., Douglas, N. J., Hames, P., & Brown, J. S. L. (2012). A randomized, placebo-controlled trial of online cognitive behavioral therapy for chronic insomnia disorder delivered via an automated media-rich web application. Sleep, 35(6), 769–781. 10.5665/sleep.1872

Henry, A. L., Miller, C. B., Emsley, R., Sheaves, B., Freeman, D., Luik, A. I., Littlewood, D. L., Saunders, K. E. A., Kanady, J. C., Carl, J. R., Davis, M. L., Kyle, S. D., & Espie, C. A. (2021). Insomnia as a mediating therapeutic target for depressive symptoms: A sub-analysis of participant data from two large randomized controlled trials of a digital sleep intervention. Journal of Sleep Research, 30(1). 10.1111/jsr.13140

Kelly, R. M., Finn, J., Healy, U., Gallen, D., Sreenan, S., McDermott, J. H., & Coogan, A. N. (2020). Greater social jetlag associates with higher HbA1c in adults with type 2 diabetes: a cross sectional study. Sleep Medicine, 66, 1–9. 10.1016/j.sleep.2019.07.023

Kind, P., Dolan, P., Gudex, C., & Williams, A. (1998). Variations in population health status: Results from a United Kingdom national questionnaire survey. British Medical Journal, 316(7133), 736–741. 10.1136/bmj.316.7133.736

Koopman, A. D. M., Rauh, S. P., Van ‘T Riet, E., Groeneveld, L., Van Der Heijden, A. A., Elders, P. J., Dekker, J. M., Nijpels, G., Beulens, J. W., & Rutters, F. (2017). The Association between Social Jetlag, the Metabolic Syndrome, and Type 2 Diabetes Mellitus in the General Population: The New Hoorn Study. Journal of Biological Rhythms, 32(4), 359–368. 10.1177/0748730417713572

Kyle, S. D., Hurry, M. E. D., Emsley, R., Marsden, A., Omlin, X., Juss, A., Spiegelhalder, K., Bisdounis, L., Luik, A. I., Espie, C. A., & Sexton, C. E. (2020). The effects of digital cognitive behavioral therapy for insomnia on cognitive function: A randomized controlled trial. Sleep, 43(9), 1–12. 10.1093/sleep/zsaa034

Lascar, N., Brown, J., Pattison, H., Barnett, A. H., Bailey, C. J., & Bellary, S. (2018). Type 2 diabetes in adolescents and young adults. In The Lancet Diabetes and Endocrinology (Vol. 6, Issue 1, pp. 69–80). Lancet Publishing Group. 10.1016/S2213-8587(17)30186-9

Leproult, R., Deliens, G., Gilson, M., & Peigneux, P. (2015). Beneficial impact of sleep extension on fasting insulin sensitivity in adults with habitual sleep restriction. Sleep, 38(5), 707–715. 10.5665/sleep.4660

Lin, C. L., Chien, W. C., Chung, C. H., & Wu, F. L. (2018). Risk of type 2 diabetes in patients with insomnia: A population-based historical cohort study. Diabetes/Metabolism Research and Reviews, 34(1). 10.1002/dmrr.2930

Lin, X., Xu, Y., Pan, X., Xu, J., Ding, Y., Sun, X., Song, X., Ren, Y., & Shan, P. F. (2020). Global, regional, and national burden and trend of diabetes in 195 countries and territories: an analysis from 1990 to 2025. Scientific Reports, 10(1). 10.1038/s41598-020-71908-9

Liu, J., Richmond, R. C., Bowden, J., Barry, C., Dashti, H. S., Daghlas, I., Lane, J. M., Jones, S. E., Wood, A. R., Frayling, T. M., Wright, A. K., Carr, M. J., Anderson, S. G., Emsley, R. A., Ray, D. W., Weedon, M. N., Saxena, R., Lawlor, D. A., & Rutter, M. K. (2022). Assessing the Causal Role of Sleep Traits on Glycated Hemoglobin: A Mendelian Randomization Study. Diabetes Care, 45(4), 772–781. 10.2337/dc21-0089

Ma, Y., Zhou, Z., Li, X., Yan, Z., Ding, K., Xiao, H., Wu, Y., Wu, T., & Chen, D. (2022). Integrative Identification of Genetic Loci Jointly Influencing Diabetes-Related Traits and Sleep Traits of Insomnia, Sleep Duration, and Chronotypes. Biomedicines, 10(2). 10.3390/biomedicines10020368

Morin, C. M. (1993). Insomnia Severity Index. PsycTESTS Dataset. 10.1037/t07115-000

Mostafa, S. A., Hanif, W., Crowe, F., Balanos, G., Nirantharakumar, K., Ellis, J. G., & Tahrani, A. A. (2025). The effect of non-pharmacological sleep interventions on glycaemic measures in adults with sleep disturbances and behaviours: A systematic review and meta-analysis. Diabetes and Vascular Disease Research, 22(1). 10.1177/14791641241307367

Nicassio, P. M., Mendlowitz, D. R., Fussell, J. J., & Petras, L. (1985). THE PHENOMENOLOGY OF THE PRE-SLEEP STATE: THE DEVELOPMENT OF THE PRE-SLEEP AROUSAL SCALE*. Behaviour Research and Therapy, 23(3), 263–271.

Ong, K. L., Stafford, L. K., McLaughlin, S. A., Boyko, E. J., Vollset, S. E., Smith, A. E., Dalton, B. E., Duprey, J., Cruz, J. A., Hagins, H., Lindstedt, P. A., Aali, A., Abate, Y. H., Abate, M. D., Abbasian, M., Abbasi-Kangevari, Z., Abbasi-Kangevari, M., Abd ElHafeez, S., Abd-Rabu, R., … Vos, T. (2023). Global, regional, and national burden of diabetes from 1990 to 2021, with projections of prevalence to 2050: a systematic analysis for the Global Burden of Disease Study 2021. The Lancet, 402(10397), 203–234. 10.1016/S0140-6736(23)01301-6

Radloff, L. S. (1977). The CES-D Scale: A Self-Report Depression Scale for Research in the General Population. APPLIED PSYCHOLOGICAL MEASUREMENT, 1(3), 385–401.

Reynolds, A. C., Dorrian, J., Liu, P. Y., van Dongen, H. P. A., Wittert, G. A., Harmer, L. J., & Banks, S. (2012). Impact of five nights of sleep restriction on glucose metabolism, leptin and testosterone in young adult men. PLoS ONE, 7(7). 10.1371/journal.pone.0041218

Ross, J. A. D., Barron, E., McGough, B., Valabhji, J., Daff, K., Irwin, J., Henley, W. E., & Murray, E. (2022). Uptake and impact of the English National Health Service digital diabetes prevention programme: observational study. BMJ Open Diabetes Research and Care, 10(3). 10.1136/bmjdrc-2021-002736

Savin, K. L., Clark, T. L., Perez-Ramirez, P., Allen, T. S., Tristão Parra, M., & Gallo, L. C. (2023). The Effect of Cognitive Behavioral Therapy for Insomnia (CBT-I) on Cardiometabolic Health Biomarkers: A Systematic Review of Randomized Controlled Trials. In Behavioral Sleep Medicine (Vol. 21, Issue 6, pp. 671–694). Taylor and Francis Ltd. 10.1080/15402002.2022.2154213

Schipper, S. B. J., Van Veen, M. M., Elders, P. J. M., Van Straten, A., Van Der Werf, Y. D., Knutson, K. L., Rutters, F., & Nl, F. R. (2021). Sleep disorders in people with type 2 diabetes and associated health outcomes: a review of the literature. Diabetologia, 64, 2367–2377. 10.1007/s00125-021-05541-0/Published

Shan, Z., Ma, H., Xie, M., Yan, P., Guo, Y., Bao, W., Rong, Y., Jackson, C. L., Hu, F. B., & Liu, L. (2015). Sleep duration and risk of type 2 diabetes: A meta-analysis of prospective studies. Diabetes Care, 38(3), 529–537. 10.2337/dc14-2073

Shen, L., Li, B. Y., Gou, W., Liang, X., Zhong, H., Xiao, C., Shi, R., Miao, Z., Yan, Y., Fu, Y., Chen, Y. M., & Zheng, J. S. (2025). Trajectories of Sleep Duration, Sleep Onset Timing, and Continuous Glucose Monitoring in Adults. JAMA Network Open, 8(3), e250114. 10.1001/jamanetworkopen.2025.0114

So-Ngern, A., Chirakalwasan, N., Saetung, S., Chanprasertyothin, S., Thakkinstian, A., & Reutrakul, S. (2019). Effects of two-week sleep extension on glucose metabolism in chronically sleep-deprived individuals. Journal of Clinical Sleep Medicine, 15(5), 711–718. 10.5664/jcsm.7758

Stamatakis, K. A., & Punjabi, N. M. (2010). Effects of sleep fragmentation on glucose metabolism in normal subjects. Chest, 137(1), 95–101. 10.1378/chest.09-0791

Sweeney, E. L., Jeromson, S., Hamilton, D. L., Brooks, N. E., & Walshe, I. H. (2017). Skeletal muscle insulin signaling and whole-body glucose metabolism following acute sleep restriction in healthy males. Physiological Reports, 5(23). 10.14814/phy2.13498

Tamm, S., Tse, K. Y. K., Hellier, J., Saunders, K. E. A., Harmer, C. J., Espie, C. A., Reid, M., & Kyle, S. D. (2025). Emotional Processing Following Digital Cognitive Behavioral Therapy for Insomnia in People with Depressive Symptoms: A Randomized Clinical Trial. JAMA Network Open, 8(2), e2461502. 10.1001/jamanetworkopen.2024.61502

Tasali, E., Wroblewski, K., Kahn, E., Kilkus, J., & Schoeller, D. A. (2022). Effect of Sleep Extension on Objectively Assessed Energy Intake Among Adults With Overweight in Real-life Settings A Randomized Clinical Trial. JAMA Internal Medicine, 182(4), 365–370. 10.1001/jamainternmed.2021.8098

The Diabetes Prevention Program (DPP) Research Group. (2002). The Diabetes Prevention Program (DPP): Description of lifestyle intervention. Diabetes Care, 25(12), 2165–2171. http://www.bsc.gwu.edu/dpp/manuals.

The EuroQoL Group. (1990). EuroQol*-a new facility for the measurement of health-related quality of life. In Health Policy.

Wang, X., Zhao, C., Feng, H., Li, G., He, L., Yang, L., Liang, Y., Tan, X., Xu, Y., Cui, R., Sun, Y., Guo, S., Zhao, G., Zhang, J., & Ai, S. (2023). Associations of Insomnia With Insulin Resistance Traits: A Cross-sectional and Mendelian Randomization Study. Journal of Clinical Endocrinology and Metabolism, 108(8), e574–e582. 10.1210/clinem/dgad089

Whicher, C. A., O’Neill, S., & Holt, R. I. G. (2020). Diabetes in the UK: 2019. Diabetic Medicine, 37(2). 10.1111/dme.14225

Wilson, S. J., Nutt, D. J., Alford, C., Argyropoulos, S. V., Baldwin, D. S., Bateson, A. N., Britton, T. C., Crowe, C., Dijk, D. J., Espie, C. A., Gringras, P., Hajak, G., Idzikowski, C., Krystal, A. D., Nash, J. R., Selsick, H., Sharpley, A. L., & Wade, A. G. (2010). British Association for Psychopharmacology consensus statement on evidence-based treatment of insomnia, parasomnias and circadian rhythm disorders. Journal of Psychopharmacology, 24(11), 1577–1600. 10.1177/0269881110379307

Wright, A. K., Huang, T., Carr, M. J., Premdayal, A. D., Saluja, S., Dashti, H. S., Anderson, S. G., Ray, D. W., Jones, S. E., Wood, A. R., Frayling, T. M., Weedon, M. N., Lane, J. M., Saxena, R., Liu, J., Bowden, J., Lawlor, D. A., Redline, S., & Rutter, M. K. (2025). Clinical utility of self-reported sleep duration and insomnia symptoms in type 2 diabetes prediction. Diabetologia. 10.1007/s00125-025-06503-6

Yuan, S., & Larsson, S. C. (2020). An atlas on risk factors for type 2 diabetes: a wide-angled Mendelian randomisation study. Diabetologia, 63(11), 2359–2371. 10.1007/s00125-020-05253-x

Zhao, S., Toniolo, S., Tang, Q.-Y., Scholcz, A., Ganse-Dumrath, A., Gendarini, C., Broulidakis, M. J., Thompson, S., Manohar, S. G., & Husain, M. (2025). OCTAL (Oxford Cognitive Testing Portal): A remote, cross-cultural cognitive assessment detects domain-specific aging and dementia. 10.1101/2025.06.23.25330153

Zhou, B., Rayner, A. W., Gregg, E. W., Sheffer, K. E., Carrillo-Larco, R. M., Bennett, J. E., Shaw, J. E., Paciorek, C. J., Singleton, R. K., Barradas Pires, A., Stevens, G. A., Danaei, G., Lhoste, V. P., Phelps, N. H., Heap, R. A., Jain, L., D’Ailhaud De Brisis, Y., Galeazzi, A., Kengne, A. P., … Ezzati, M. (2024). Worldwide trends in diabetes prevalence and treatment from 1990 to 2022: a pooled analysis of 1108 population-representative studies with 141 million participants. The Lancet, 404(10467), 2077–2093. 10.1016/S0140-6736(24)02317-1

Zhu, B., Shi, C., Park, C. G., Zhao, X., & Reutrakul, S. (2019). Effects of sleep restriction on metabolism-related parameters in healthy adults: A comprehensive review and metaanalysis of randomized controlled trials. In Sleep Medicine Reviews (Vol. 45, pp. 18–30). W.B. Saunders Ltd. 10.1016/j.smrv.2019.02.002

Zuraikat, F. M., Laferrère, B., Cheng, B., Scaccia, S. E., Cui, Z., Aggarwal, B., Jelic, S., & St-Onge, M. P. (2024). Chronic insufficient sleep in women impairs insulin sensitivity independent of adiposity changes: Results of a randomized trial. Diabetes Care, 47(1), 117–125. 10.2337/dc23-1156

