## Supplementary tables and figures for "Sleep improvement for metabolic health: A feasibility trial of a digital sleep treatment in people with insomnia and non-diabetic hyperglycaemia"

**Supplementary Table 1**

|  | **Pre-intervention, mean (SD)** | **Post-intervention, mean (SD)** | **Mean change score (95% C.I)** | **Cohen's d**  **(95% C.I)** |
| --- | --- | --- | --- | --- |
| **Identification Time 1 item** | 1.6 (0.4) | 1.6 (0.4) | 0.1 (-0.1 to 0.2) | 0.3 (-0.2 to 0.7) |
| **Identification Time 3 item** | 3.2 (1.0) | 3.0 (0.9) | -0.1 (-0.6 to 0.3) | -0.1 (-0.6 to 0.3) |
| **Localisation Time 1 item** | 4.5 (0.9) | 4.2 (1.1) | -0.2 (-0.5 to 0.2) | -0.2 (-0.6 to 0.2) |
| **Localisation Time 3 item** | 6.7 (1.8) | 6.2 (1.9) | -0.3 (-0.9 to 0.2) | -0.3 (-0.7 to 0.2) |
| **Proportion Correct 1 item** | 1.0 (0.0) | 1.0 (0.0) | -0.0 (-0.0 to 0.0) | -0.2 (-0.6 to 0.3) |
| **Proportion Correct 3 item** | 0.9 (0.1) | 0.9 (0.1) | 0.0 (-0.0 to 0.1) | 0.2 (-0.2 to 0.7) |

Supplemental Table 1: Pre-intervention and post-intervention means, standard deviations (SD), pre-to-post change scores, and effect sizes (Cohen’s d) for the visual short-term working memory task. 95% confidence intervals (C.I) are presented for the mean change and the effect size.

**Supplementary Table 2**

|  | **Pre-intervention, mean (SD)** | **Post-intervention, mean (SD)** | **Mean change score (95% C.I)** | **Cohen's d**  **(95% C.I)** |
| --- | --- | --- | --- | --- |
| **Number of days CGM worn out of 7** | 6.6 (0.5) | 6.7 (0.5) | 0.1 (-0.2 to 0.3) | 0.1 (-0.4 to 0.6) |
| **Mean sensor glucose level** | 6.6 (0.5) | 6.6 (0.8) | 0.0 (-0.4 to 0.4) | 0.0 (-0.4 to 0.5) |
| **Mean sensor glucose level during "sleep"** | 6.3 (0.6) | 6.2 (0.8) | 0.0 (-0.3 to 0.4) | 0.1 (-0.4 to 0.5) |
| **Mean sensor glucose level during "wake”** | 6.7 (0.5) | 6.8 (0.9) | 0.0 (-0.4 to 0.5) | 0.0 (-0.4 to 0.5) |
| **Coefficient of variation for glucose (CV)** | 19.0 (2.8) | 19.9 (4.0) | 0.4 (-1.3 to 2.0) | 0.1 (-0.4 to 0.6) |
| **Coefficient of variation for glucose (CV) during "sleep"** | 10.1 (3.1) | 10.9 (3.4) | 0.9 (-0.7 to 2.5) | 0.3 (-0.2 to 0.7) |
| **Coefficient of variation for glucose (CV) during "wake"** | 20.3 (2.8) | 21.0 (4.3) | 0.3 (-1.4 to 2.1) | 0.1 (-0.4 to 0.6) |
| **Percentage of time in range (3.9-10 mmol/L)** | 97.2 (1.6) | 95.1 (4.7) | -1.6 (-3.9 to 0.6) | -0.3 (-0.8 to 0.1) |
| **Percentage of time in range (3.9-10 mmol/L) during "sleep"** | 99.2 (1.7) | 98.5 (3.0) | -0.7 (-2.2 to 0.8) | -0.2 (-0.7 to 0.2) |
| **Percentage of time in range (3.9-10 mmol/L) during "wake"** | 96.5 (2.0) | 94.0 (6.0) | -2.0 (-4.8 to 0.8) | -0.3 (-0.8 to 0.1) |

Supplemental Table 2: Pre-intervention and post-intervention means, standard deviations (SD), pre-to-post change scores, and effect sizes (Cohen’s d) for the continuous glucose monitor. Outcomes were averaged over the sensor days worn. 95% confidence intervals (C.I) are presented for the mean change and the effect size. “Sleep” time was defined between the hours of 00:00 and 06:00 whereas “wake” time was defined as 06:01 to 23:59. Glucose levels are recorded in mmol/L

**Supplementary Table 3**

|  | **Pre-intervention** | **Post-intervention** |
| --- | --- | --- |
| **Work-related accident or injury in the last two months, n(%)** | 0 (0.0%) | 0 (0.0%) |
| **Motor vehicle accident in the last two months, n(%)** | 0 (0.0%) | 0 (0.0%) |
| **Near miss incident in the last two months, n(%)** | 0 (0.0%) | 0 (0.0%) |
| **Times fallen asleep driving in the last two months, mean(SD)** | 0.0 (0.2) | 0 (0.0) |
| **Number of falls last in the last two months, mean(SD)** | 0.1 (0.3) | 0.2 (0.5) |

Supplemental Table 3: Pre-intervention and post-intervention n, means, and standard deviations (SD), pre-to-post for the pre-specified trial adverse events.

**Supplementary Figure 1**

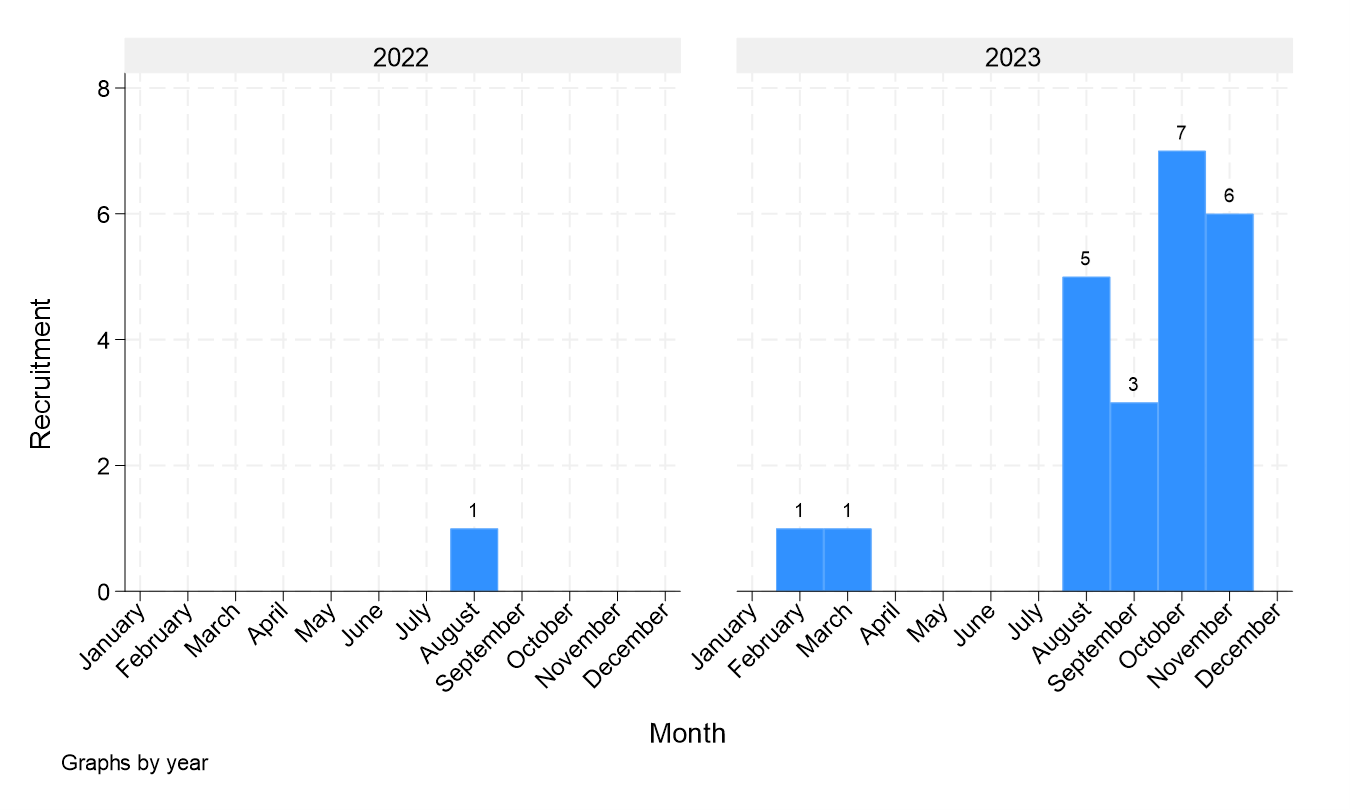

Supplemental Figure 1: Bar graph showing enrolment rates per month. The study ran from August 2022 to November 2023.

**Supplementary Figure 2**

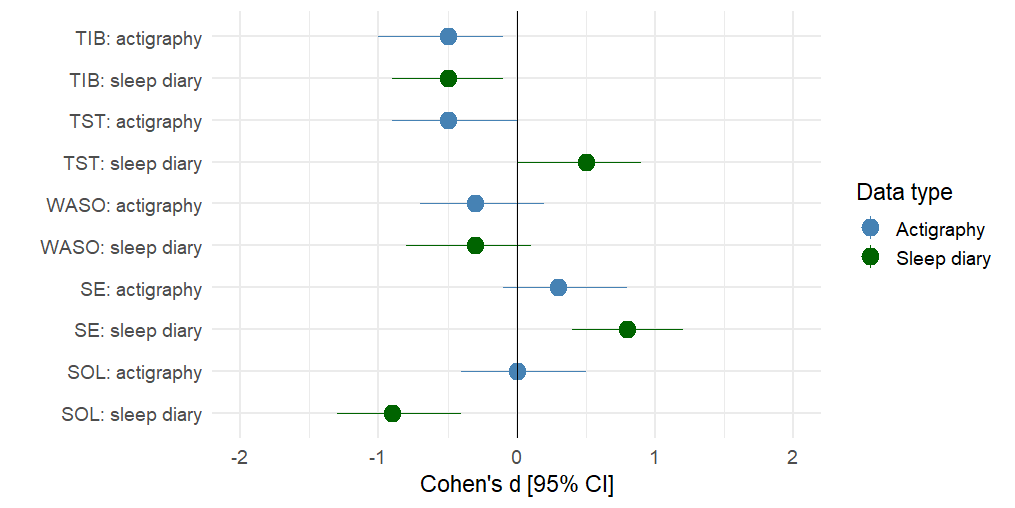

Supplemental Figure 2: Forest plot showing the 95% confidence intervals of the effect sizes for the matched actigraphic and sleep diary variables. TIB: Time In Bed. TST: Total Sleep Time. WASO: Wake After Sleep Onset. SE: Sleep Efficiency. SOL: Sleep Onset Latency
